# Alcohol use disorder and epileptogenesis in primary malignant brain tumors: temporal and tumor grade associations in a nationwide EHR cohort study

**DOI:** 10.64898/2025.12.11.25342079

**Authors:** Kevin Hoffman, Malay Shah, James Madsen, Damisi Akinpelu, Jochen Meyer

## Abstract

**Background:** Primary malignant brain tumors (PMBTs), including glioblastomas (GBM) and low-grade gliomas (LGGs), frequently cause seizures, which worsen patient morbidity and quality of life. Alcohol use disorder (AUD) is a known risk factor for seizures in the general population, but its role in PMBT-associated seizures remains poorly understood.

**Objective:** To evaluate the association between AUD and seizure prevalence in PMBT patients, and to examine whether this relationship varies by age, sex, race/ethnicity, and tumor grade.

**Methods:** We conducted a retrospective cohort study using the Cosmos EHR database (2015-2025), identifying 244,955 adult patients with PMBTs. Seizure/epilepsy and AUD diagnoses were ascertained via ICD-10 codes. Associations were assessed using chi-square tests and logistic regression (adjusting for age, sex, and race). Temporal sequencing of AUD and seizure diagnoses was analyzed in six-month intervals. Subgroup analyses were performed for glioma grade (low vs. high).

**Results:** PMBT patients with seizures were nearly twice as likely to have AUD compared to those without (OR = 1.90, 95% CI 1.83-1.97, p < 0.001). After adjusting for demographics, AUD remained significantly associated with seizure risk (OR = 1.37, 95% CI 1.24-1.51, p < 0.001). This association was strongest in younger patients and present across all sexes and racial groups. Temporal analyses indicated that AUD partly preceded seizure onset. Among patients with available histologic data, alcohol-using low-grade patients exhibited a markedly higher seizure prevalence compared to high-grade patients (43.8% vs. 12.1%, p < 0.001).

**Conclusions:** AUD is potentially associated with increased seizure risk in PMBT patients, with a stronger effect in younger individuals and low-grade tumors. These findings suggest that alcohol-related hyperexcitability may compound tumor-associated epileptogenesis, highlighting AUD as a potentially modifiable risk factor. Prospective studies are warranted to confirm these relationships and to evaluate interventions targeting alcohol use for seizure mitigation.

**Key Points:**

- PMBT patients with seizures had higher AUD prevalence (OR = 1.90, 95% CI 1.83-1.97).
- Models controlling for demographic variables confirmed increased seizure risk persisted in the AUD population (OR = 1.37, 95% CI 1.24-1.51).
- Association strongest in younger patients, unchanged across sex and race groups.
- Seizures were more common in AUD low-grade gliomas vs high-grade (43.8% vs 12.1%).
- Temporal data suggest AUD partly preceded seizure onset in PMBT patients.

## Introduction

Primary malignant brain tumors (PMBTs) are the most aggressive and clinically significant neoplasms of the central nervous system in adults. The majority are gliomas, with approximately 49% being glioblastoma multiforme (GBM), or glioma grade 4, and 30% being lower-grade gliomas, demonstrating the spectrum of tumor aggressiveness within PMBTs. Besides symptoms like headaches, cognitive decline, and neurologic deficits, seizures are a major comorbidity affecting patients (Schaff & Mellinghoff, 2023). Between 30-45% of patients present with seizures at diagnosis, and an additional 10-30% develop them during the disease course (Feyissa et al., 2023). Moreover, seizures are more common in low-grade gliomas (LGGs) than in high-grade gliomas (HGGs). This may be because the slow growth of low-grade tumors allows time for the brain to develop the local and/or distant cellular changes that promote seizures (van Breemen, 2007). While their effect on overall survival remains uncertain, seizures are consistently linked to poor quality of life and increased morbidity in PMBT patients (Dobran et al., 2018; Krajewski et al., 2021). Additionally, recent molecular and clinical studies have shown that tumor-neuron interactions, synaptic remodeling, and the hyperexcitable microenvironments contribute to the elevated seizure risk in PMBT patients, highlighting the complex mechanisms of epileptogenesis associated with these tumors (Huang-Hobbs et al., 2023; Soeung et al., 2024; Meyer et al., 2024).

Alcohol use disorder (AUD) has been identified as a risk factor for seizures, with an overall twofold higher risk of unprovoked seizures and epilepsy among those who consume alcohol compared to those who do not (Samokhvalov et al., 2010). AUD affects neuronal excitability by enhancing glutamatergic signaling and suppressing GABAergic inhibition, and this effect is dose dependent (Becker & Mulholland, 2014). Consequently, episodes of alcohol withdrawal lead to hyperexcitable states due to this compensatory neuroexcitation (Hillbom et al., 2003; Ng et al., 1988). Despite these established associations, the relationship between AUD and seizure prevalence specifically in PMBT patients remains poorly understood. Seizures in brain tumor patients are often attributed to the tumor’s structural and physiological effects. There is limited attention given to modifiable factors such as alcohol use (Walsh et al., 2017).

Therefore, understanding the role of AUD in PMBT patients is important, since addressing modifiable factors can improve both quality of life and seizure management. Furthermore, seizure activity and neuronal hyperexcitability can affect tumor growth and invasion (Goethe et al., 2023). This suggests that behavioral interventions targeting alcohol use could have benefits beyond seizure control, potentially modulating tumor progression.

## Methods

### Data Source

We conducted a retrospective cohort study of patient diagnoses over a ten-year period (June 26, 2015 - June 25, 2025) using the Cosmos research platform (Epic Systems Corporation; Verona, WI). Cosmos aggregates electronic health record (EHR) data submitted voluntarily by health systems for the purposes of research. At the time of submission, the Cosmos data set comprised 296 million patients and 1700 hospitals. Epic centrally de-duplicated and anonymized the patient records before research teams accessed data.

### Study Population

We used ICD-10 codes to query Cosmos for adult patient diagnoses from June 26, 2015, to June 25, 2025. As the Cosmos database uses the ICD-10-CM system, it does not include extensive data on histological record or molecular subtype. Thus, diagnoses were identified via ICD-10 codes for malignant neoplasm of the brain (ICD-10-CM C71.*), which entails tumors of several cell types (glioblastoma, astrocytoma, oligodendrocytoma, ependymoma). To further specify tumors of interest with the limitation of Cosmos’ classification system, we excluded patients with benign brain tumors (ICD-10-CM D33.*). The cohort was further stratified by presence or absence of seizure or epilepsy diagnoses (ICD-10-CM G40.*, R56.*) and alcohol-related disorders (ICD-10-CM F10.*) following similar cohort designation in prior literature (Calonge et al., 2025). For analyses involving glioma grade, patients with documented histologic classification as low-grade or high-grade glioma were selected, using available Histologic Grade records linked in the EHR. Low-grade was defined as Grade 1/I, Grade 2/II, and Low Grade, while High-grade was defined as Grade 3/III, Grade 4/IV, and High-grade. The study period for this subgroup was extended from 2010 to 2025 to increase the sample size due to paucity of histological records.

We analyzed the queried variables based on exposure of alcohol-related disorder diagnoses (ICD-10-CM F10.*) related to the outcome variable of seizure or epilepsy diagnoses (ICD-10-CM G40.* or R56.*). Covariate analyses were performed to evaluate modifiers of the outcome variable. We designated demographic variables of interest in following prior literature that examined drug exposure and clinical diagnose outcomes using the Cosmos data base (Shalaby et al., 2025). We examined age (categorized as 18-40, 40-65, and 65+ years), sex (male/female), and race/ethnicity (White, Black or African American, Asian, American Indian/Alaska Native + Native Hawaiian/Pacific Islander [AI/AN + NH/PI], and Other).

### Temporal Analysis

To investigate the temporal relationship between alcohol use and seizure/epilepsy diagnoses, we identified malignant brain tumor patients with incident (new) diagnoses of alcohol-related disorders (ICD-10-CM F10.*) and seizures/epilepsy (ICD-10-CM G40.* or R56.*) and analyzed diagnosis sequencing over a 102-month follow-up. Beyond 102-months, there was insufficient statistical power for Cosmos’ anonymization (<10 records). Time intervals were defined in 6-month increments, and counts of patients with alcohol diagnosis preceding seizure diagnosis (AUD prior to Epilepsy) and vice versa (Epilepsy prior to AUD) were calculated.

### Statistical Analysis

We performed chi-square tests to evaluate associations between alcohol use and seizure/epilepsy diagnoses overall and within stratified groups (age, sex, race/ethnicity). Odds ratios (OR) with 95% confidence intervals (CI) quantified effect sizes. Further, we assessed the overall association and homogeneity, controlling for each of the three stratification variables (age, sex, race/ethnicity) using Mantel-Haenszel common odds ratios and Breslow-Day tests (Mantel and Haenszel, 1959 & Breslow, 1980). We constructed a linear regression model to estimate the association between alcohol use and seizure-use in glioma patients adjusting for age, sex, and race. Temporal association between (Alcohol → Epilepsy) and (Epilepsy → Alcohol) was assessed via chi-square and Fisher’s exact test at each time interval. Odds ratios for alcohol use preceding seizure compared to the reverse were calculated with 95% CIs. Lastly, we applied Fisher’s exact test to examine the seizure prevalence between low-grade and high-grade glioma patients with alcohol use. All statistical tests were two-sided, and p-values < 0.05 were considered statistically significant. Analyses were conducted using Python 3.13.5.

## Results

From June 26, 2015 to June 25, 2025, we identified 244,955 adult patients in the Cosmos EHR database with a diagnosis of malignant neoplasm of the brain (ICD-10-CM C71.*), excluding those with benign tumors (ICD-10-CM D33.*). Patients were stratified by the presence of epilepsy or seizure diagnoses (ICD-10-CM G40.*, R56.*) and alcohol-related disorders (ICD-10-CM F10.*).

**Table 1:** Demographics table with counts and percentage proportion from the total population of PMBT patients from 2015-2025.

| Baseline characteristic |  |  |
| --- | --- | --- |
|  | *n* | % |
| Gender |  |  |
| Female | 127,902 | 52.3 |
| Male | 116,639 | 47.7 |
| Age |  |  |
| 18-40 | 36,817 | 16.1 |
| 40-65 | 86,889 | 38.0 |
| 65+ | 104,669 | 45.8 |
| Race/Ethnicity |  |  |
| White | 201,919 | 83.3 |
| Black or African<br>American | 23,309 | 9.6 |
| Asian | 7,709 | 3.2 |
| AI/AN + NH/PI | 3,315 | 1.4 |
| Other | 6,128 | 2.5 |

Among the 244,955 PMBT patients identified by our criteria. The total population of each demographic variable was slightly less than 244,955, likely due to slight gaps in patient records for recording the relevant variables. With nearly 45.8% of the population older than 65, our study population skews towards older patients. Furthermore, the study population is predominantly white (83.3%). Though we later control for these variables through logistic regression, these patterns should be considered when interpreting associations between alcohol and seizure risk in PMBT populations.

**Figure 1.**
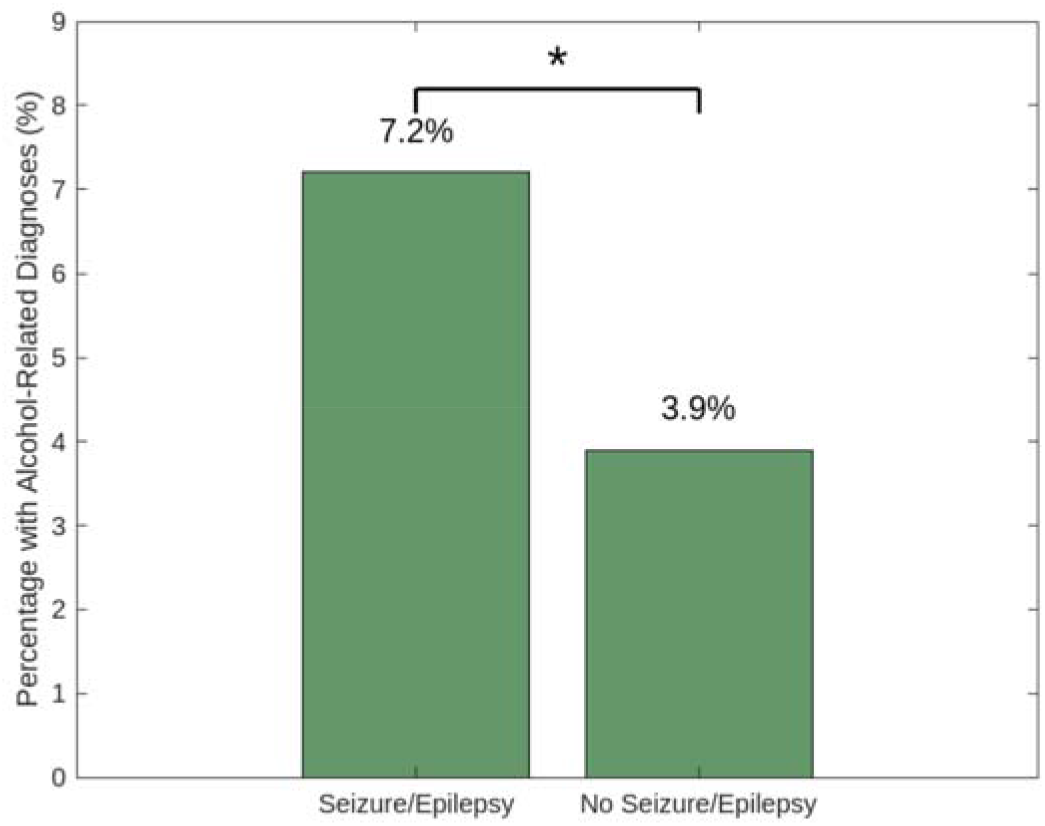
PMBT patients with seizures/epilepsy were nearly twice as likely to have alcohol-related diagnoses as those without (Chi-square, OR = 1.90; 95% CI: 1.83-1.97; *p* < 0.0001).

Among the 88,131 patients with seizures or epilepsy, 6,377 (7.2%) also had alcohol-related diagnoses. In contrast, among the 156,824 patients without seizure or epilepsy diagnoses, 6,190 (3.9%) had alcohol-related diagnoses. This difference was statistically significant via the chisquare test. This suggests a strong and statistically significant association between alcohol use and the presence of seizure or epilepsy symptoms among patients with malignant brain tumors. This compares to a prevalence of around 0.5% to 1% of the general population who are diagnosed with epilepsy and around 5% of healthy individuals diagnosed with AUD who develop epilepsy (Fiest et al., 2017 & Bakken et al., 2014).

### Demographic Analysis

We assessed whether the association between alcohol-use and seizures varied across demographic groups. We evaluated the relationship between alcohol use and seizure/epilepsy diagnoses among PMBT patients using generalized linear logistic regression models, adjusting for age, sex, and race.

**Figure 2.**
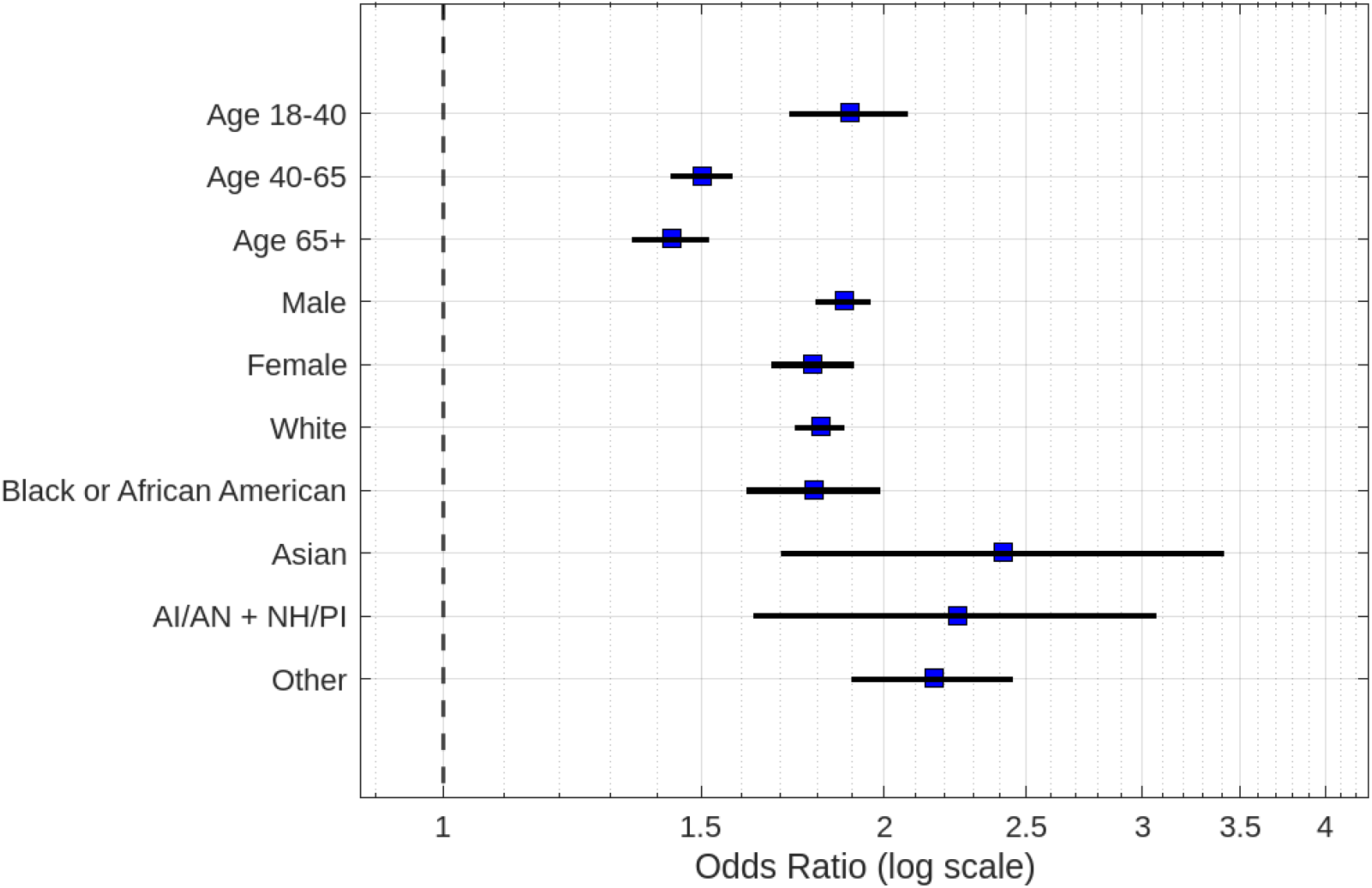
Logistic regression odds ratio of seizure risk within each demographic variable based on alcohol-use (CI 95% and p < 0.05).

Alcohol use was associated with increased seizure prevalence across all age groups, though the magnitude declined with age (Logistic regression, 18-40 years: OR 1.89, 95% CI 1.72-2.08; 40-65 years: OR 1.50, 95% CI 1.43-1.58; 65+ years: OR 1.43, 95% CI 1.35-1.52; all p < 0.0001). This suggests that age may modify the relationship between alcohol use and seizure prevalence in PMBT patients.

Both male and female patients who used alcohol had significantly higher odds of seizures compared to their non-using counterparts in males (OR 1.88, 95% CI 1.80-1.96) and in females (OR 1.79, 95% CI 1.67-1.91), with p < 0.0001 for both groups. Though there was a small increase in prevalence in males than in females, it is non-significant, so sex may not modify the relationship between alcohol use and seizure prevalence.

Across all racial demographics analyzed, alcohol use was significantly associated with increased odds of seizures. Odds ratios ranged from 1.79-2.41, with the highest relative risk observed in Asian patients (OR 2.41, 95% CI 1.70-3.41), followed by AI/AN + NH/PI (OR 2.24, 95% CI 1.63-3.07) and ‘Other’ (OR 2.16, 95% CI 1.90-2.45). White (OR 1.81, 95% CI 1.74-1.88) and Black or African American (OR 1.79, 95% CI 1.61-1.99) all p < 0.0001. However, with significant variation within each category, it is unclear whether specific race/ethnicity may act as a modifier in the relationship between alcohol use and seizure prevalence.

**Figure 3.**
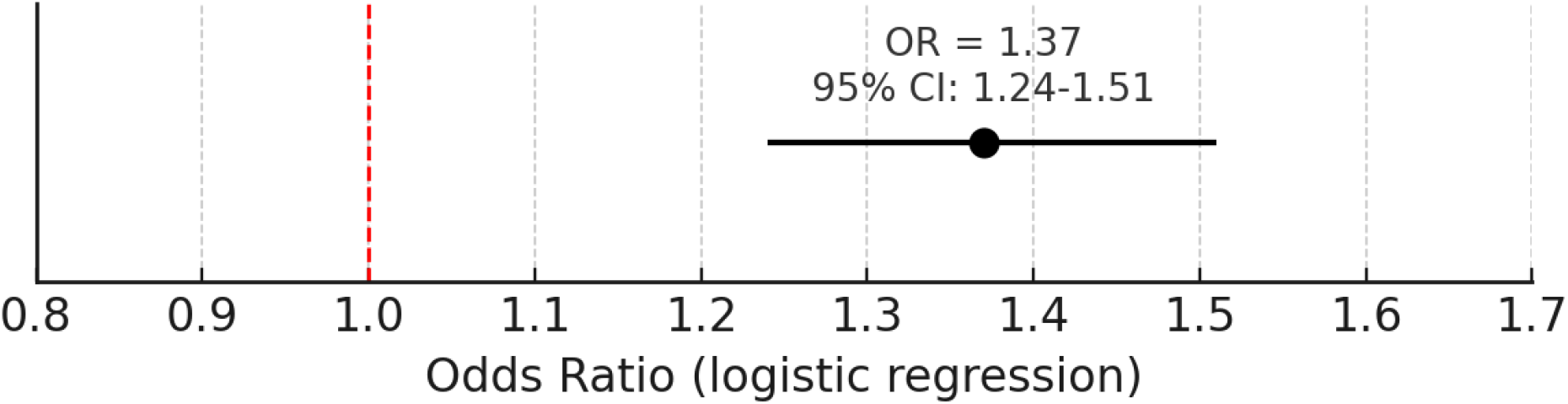
Association between alcohol use and seizure/epilepsy diagnosis. Logistic regression demonstrated that alcohol use was associated with significantly increased odds of seizure or epilepsy diagnosis (OR = 1.37, 95% CI: 1.24-1.51, p < 0.001), adjusting for age, sex, and race.

Overall logistic regression analysis demonstrated that alcohol use was significantly associated with increased odds of seizure or epilepsy diagnosis (OR = 1.37, 95% CI: 1.24-1.51, p < 0.001), even after adjusting for demographic factors including age, sex, and race.

### Temporal Analysis

To investigate the temporal association between alcohol use and seizure or epilepsy diagnoses in glioblastoma (GBM) patients, we analyzed new diagnoses (incident cases) of alcohol-related disorders (ICD F10.*) and seizure/epilepsy (ICD G40.*, R56.*) over a 102-month follow-up period.

**Figure 4.**
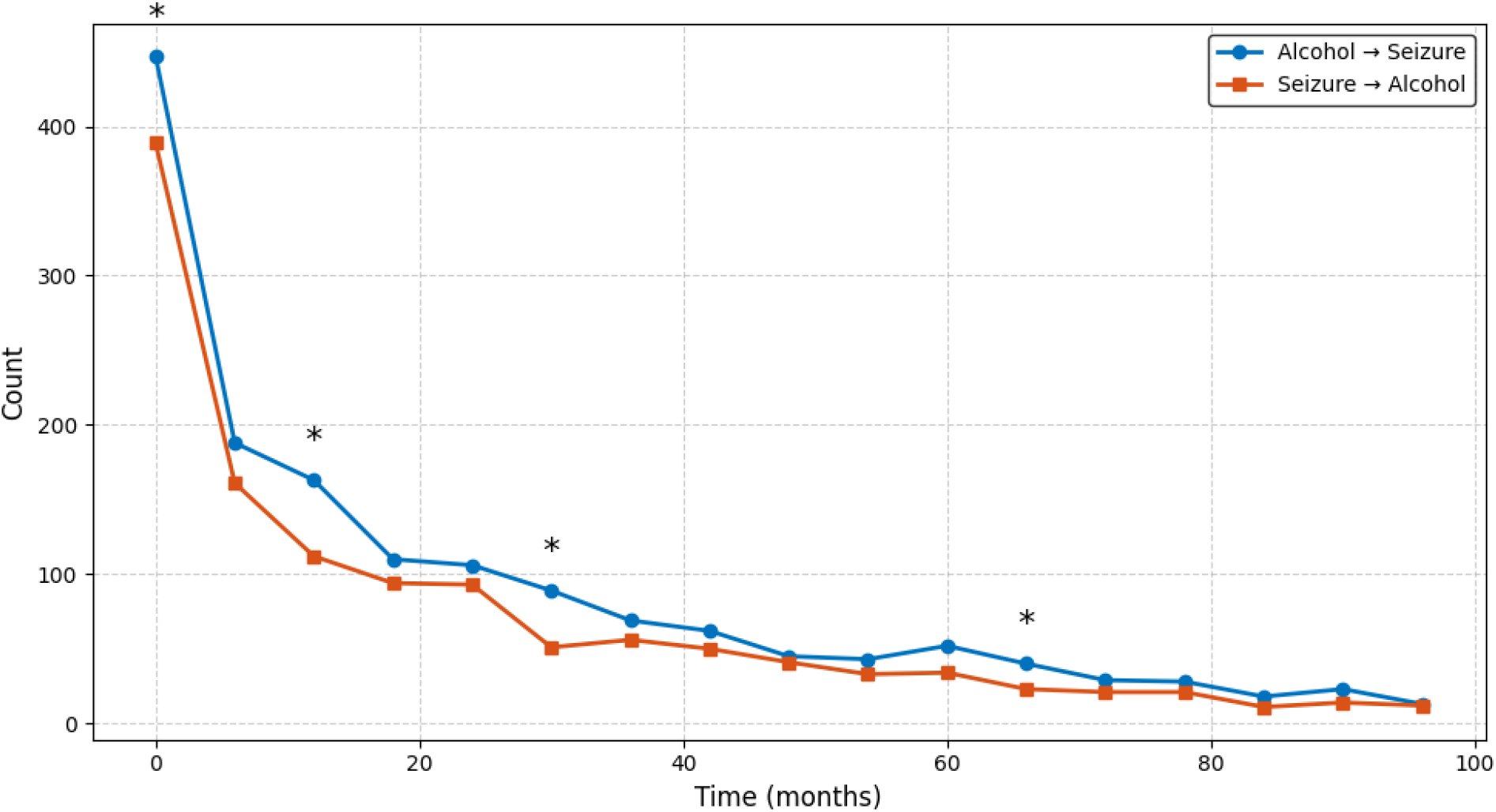
Temporal count of new epilepsy and alcohol diagnoses anchored on months after first alcohol or epilepsy diagnosis respectively. Line plots show the number of patients with AUD preceding seizures/epilepsy diagnosis and seizures/epilepsy preceding AUD diagnosis over time in months. Counts are plotted in 6-month intervals up to 102 months. Asterisks (*) indicate timepoints where group differences were statistically significant via Chi-square tests (*p* < 0.05).

At every interval, more patients were diagnosed with AUD prior to seizure onset than the reverse, though this relationship is only significant at a few time points including 0-6 months (OR 1.16, 95% CI 1.01-1.34, p = 0.041), 12-18 months (OR 1.47, 95% CI 1.15-1.87, p = 0.002), 30-36 months (OR 1.76, 95% CI 1.24-2.48, p = 0.002), and 66-72 months (OR 1.74, 95% CI 1.04-2.92, p = 0.043). While most intervals did not reach statistical significance, the overall pattern consistently indicated that alcohol use often preceded seizure diagnosis, with odds ratios generally greater than 1.0. However, without adjacent timepoints reaching statistical significance, it is difficult to conclude a precise temporal pattern.

### Tumor Grade Analyses

To further investigate the relationship between glioma grade, seizures/epilepsy, and alcohol use, we analyzed patients with malignant brain neoplasms (ICD-10: C71.*) stratified by histologic tumor grade (low-grade vs. high-grade) and presence of alcohol use disorder (ICD-10: F10.*) using Cosmos EHR data from 2010 to 2025. The dataset timeline was expanded due few HER records of pathology reports leading to privacy-related data suppression, leaving low-grade glioma seizure/epilepsy counts reported as <10 from 2015-2025.

**Figure 5.**
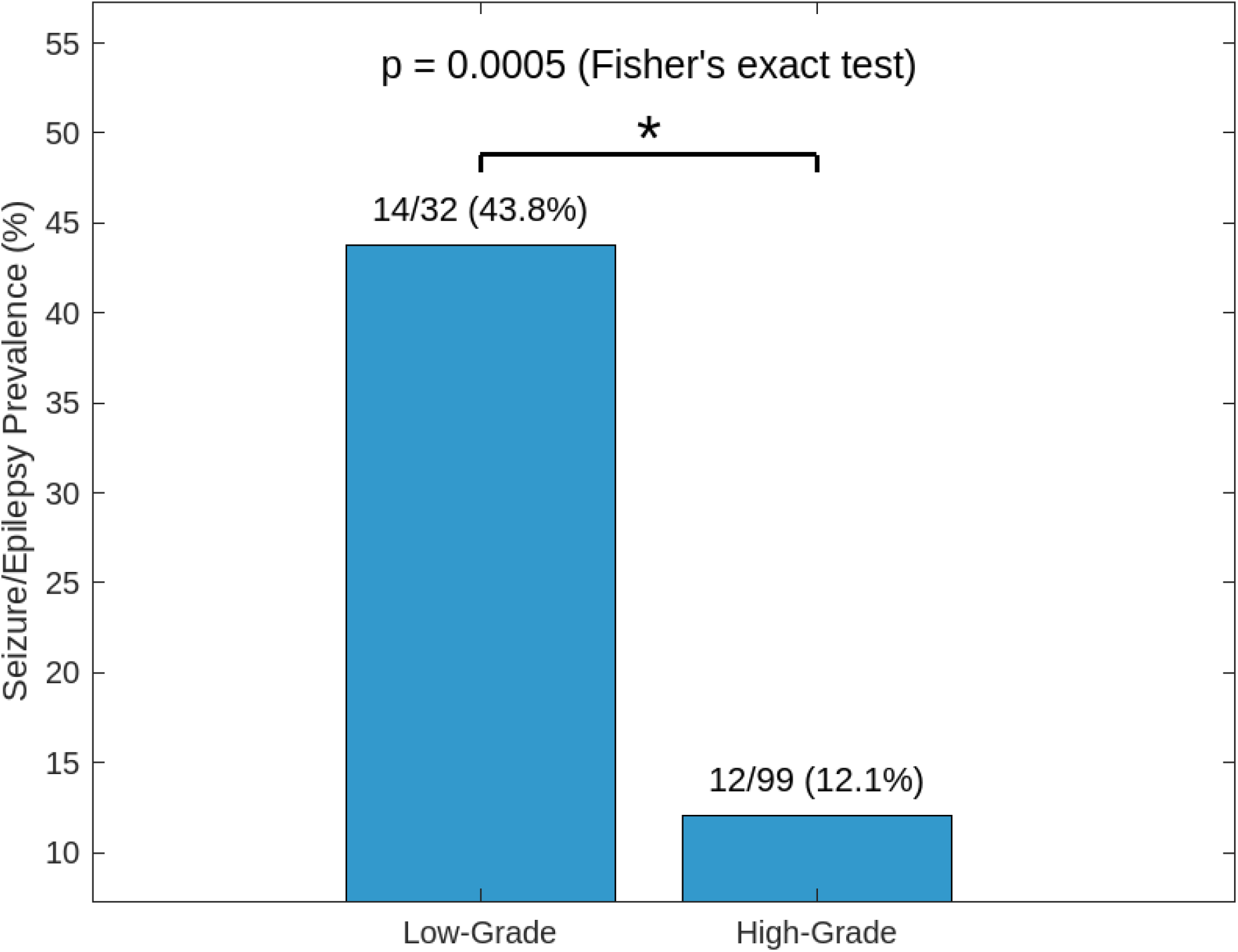
Seizure prevalence among AUD PMBT patients by tumor grade. Low grade 32 alcohol users, 14 seizure/epilepsy cases, 43.8% with seizures. High grade 99 alcohol users, 12 with seizures, 12.1%. Fisher’s exact test (OR = 5.64; p = 0.0005).

Though limited by small sample sizes and data suppression thresholds, seizure rates were statistically higher in the low-grade glioma patient population than high-grade glioma patients. However, these findings are very limited by the available histological data, so further exploration may help to elicit whether alcohol use affects seizure/epilepsy prevalence in low-grade tumor populations compared to high-grade tumor populations.

## Discussion

In this multi-institutional retrospective cohort study of patients with primary malignant brain tumors (PMBT), alcohol-related disorders were associated with significantly increased odds of seizures and epilepsy (unadjusted OR = 1.90; adjusted OR = 1.37, 9% CI 1.24-1.51). This effect persisted after adjustment for age, sex, and race/ethnicity. While the strength of association varied by demographic subgroup, it was more pronounced in younger patients (18-40), suggesting that age-related patterns of alcohol use may influence seizure vulnerability.

Temporal analyses showed that alcohol use more often preceded seizure onset than the reverse, with this directional pattern appearing early and persisting throughout follow-up. These findings support alcohol as a potential contributor to seizure pathogenesis, rather than seizures being the primary driver of alcohol use.

Stratification by tumor grade revealed a substantially higher prevalence of seizures among alcohol-using patients with low-grade gliomas compared with those with high-grade tumors. This aligns with prior literature noting the greater epileptogenic potential of low-grade gliomas and suggests that alcohol use compounds seizure risk in a way consistent with broader glioma-associated epileptogenesis.

Prior epidemiologic studies in the general population have consistently shown that alcohol use disorder confers nearly a twofold increased risk of epilepsy and unprovoked seizures, with risk particularly elevated during withdrawal states. Mechanistically, heavy alcohol consumption lowers seizure threshold through processes such as neurotoxicity, glutamatergic upregulation, impaired GABAergic inhibition, thiamine deficiency, and withdrawal effects (Samokhvalov, et al., 2010). These mechanisms may intensify tumor-associated synaptic remodeling and excitatory microenvironments in gliomas (Soeung et al., 2024, Meyer et al., 2024). Our findings extend this body of evidence by demonstrating a similar association in the PMBT population, suggesting that alcohol-related hyperexcitability can amplify tumor-driven epileptogenesis.

Historically, tumor-related epilepsy and alcohol-related epilepsy have been treated as distinct entities. Our study bridges these domains by showing that alcohol use is itself a risk factor for seizures in patients with malignant brain tumors. This integrative view is novel and highlights alcohol as a compounding factor in an already vulnerable population. In glioblastoma and other PMBTs, seizures are among the most frequent presenting symptoms, reported in 30-45% of cases. Although some studies describe epilepsy in gliomas as a favorable prognostic marker, others (e.g., Dobran et al.) have found no independent prognostic value after accounting for age, tumor location, and extent of resection. However, other studies have found age itself playing a modulatory role in developing seizures in glioma patients (Xiao et al., 2025). Our study suggests that alcohol use in younger patients may explain this phenomenon. Indeed, our results add nuance to this debate by suggesting that alcohol use may represent a modifiable risk factor that operates independently of tumor biology.

This study has several limitations. The retrospective design precludes causal inference, reliance on ICD-10 coding introduces risk of misclassification for both alcohol use and seizure diagnoses, and incomplete histopathologic reporting of tumor grade reduced subgroup sample sizes (n = 32 low-grade, n = 99 high-grade), limiting statistical power. However, prior studies have been limited by less representative datasets, while the Cosmos database provides a large, diverse, multi-institutional population, and we applied standardized query definitions and analytic methods to minimize potential biases. Prospective, well-characterized studies are needed to validate these associations, clarify biological pathways linking alcohol use and seizure risk, and evaluate whether alcohol reduction interventions can mitigate seizure burden in this patient group.

Finally, prior research (e.g., Samokhvalov et al., 2010) has demonstrated a dose-dependent relationship between alcohol consumption and seizure occurrence. Our study defined exposure by presence of alcohol-related disorder, without stratifying by level of use. Future investigations should quantify alcohol intake and explore interactions with age, as younger patients-who often engage in heavy or binge drinking-may be at disproportionately higher risk. Disentangling tumor-related seizure risk by both age and alcohol consumption would add clarity, as most large glioma-epilepsy cohorts have not systematically assessed these combined effects.

## Conclusion

In this large EHR-based cohort, AUD was significantly associated with increased seizure risk in PMBT patients, particularly in younger individuals and those with low-grade tumors. These findings suggest that AUD may contribute to tumor-related epileptogenesis. This may indicate alcohol use as a potentially modifiable risk factor. Prospective studies are needed to confirm these associations and evaluate targeted interventions.

## Attributions

Data used in this study came from Epic Cosmos, a dataset created in collaboration with a community of Epic health systems representing more than 300 million patient records from over 1762 hospitals and 40700 clinics as of August 2025. The community represents patients from all 50 states, D.C., Canada, Lebanon, and Saudi Arabia.

## Data Availability

All data produced in the present study are available upon reasonable request to the authors

## Acknowledgments

The authors thank Dr. Zakir Mridha for their assistance.

## References

1. Schaff, L. R., & Mellinghoff, I. K. (2023). Glioblastoma and Other Primary Brain Malignancies in Adults: A Review. JAMA, 329(7), 574–587. 10.1001/jama.2023.0023

2. Feyissa, A. M., Sanchez-Boluarte, S. S., Moniz-Garcia, D., Chaichana, K. L., Sherman, W. J., Freund, B. E., Tatum, W. O., Middlebrooks, E. H., Sirven, J. I., & Quinones-Hinojosa, A. (2023). Risk factors for preoperative and postoperative seizures in patients with glioblastoma according to the 2021 World Health Organization classification. Seizure, 112, 26–31. 10.1016/j.seizure.2023.09.013

3. van Breemen, M. S., Wilms, E. B., & Vecht, C. J. (2007). Epilepsy in patients with brain tumours: epidemiology, mechanisms, and management. The Lancet. Neurology, 6(5), 421–430. 10.1016/S1474-4422(07)70103-5

4. Dobran, M., Nasi, D., Chiriatti, S., Gladi, M., Somma, L. D., Iacoangeli, M., & Scerrati, M. (2018). Prognostic Factors in Glioblastoma: Is There a Role for Epilepsy?. Neurologia medico-chirurgica, 58(3), 110–115. 10.2176/nmc.oa.2017-0167

5. Krajewski, S., Wójcik, M., Harat, M., & Furtak, J. (2021). Influence of Epilepsy on the Quality of Life of Patients with Brain Tumors. International journal of environmental research and public health, 18(12), 6390. 10.3390/ijerph18126390

6. Huang-Hobbs, E., Cheng, Y. T., Ko, Y., Luna-Figueroa, E., Lozzi, B., Taylor, K. R., McDonald, M., He, P., Chen, H. C., Yang, Y., Maleki, E., Lee, Z. F., Murali, S., Williamson, M. R., Choi, D., Curry, R., Bayley, J., Woo, J., Jalali, A., Monje, M., … Deneen, B. (2023). Remote neuronal activity drives glioma progression through SEMA4F. Nature, 619(7971), 844–850. 10.1038/s41586-023-06267-2

7. Soeung, V., Puchalski, R. B., & Noebels, J. L. (2024). The complex molecular epileptogenesis landscape of glioblastoma. Cell reports. Medicine, 5(8), 101691. 10.1016/j.xcrm.2024.101691

8. Meyer, J., Yu, K., Luna-Figueroa, E., Deneen, B., & Noebels, J. (2024). Glioblastoma disrupts cortical network activity at multiple spatial and temporal scales. Nature communications, 15(1), 4503. 10.1038/s41467-024-48757-5

9. Samokhvalov, A. V., Irving, H., Mohapatra, S., & Rehm, J. (2010). Alcohol consumption, unprovoked seizures, and epilepsy: a systematic review and meta-analysis. Epilepsia, 51(7), 1177–1184. 10.1111/j.1528-1167.2009.02426.x

10. Hillbom, M., Pieninkeroinen, I., & Leone, M. (2003). Seizures in alcohol-dependent patients: epidemiology, pathophysiology and management. CNS drugs, 17(14), 1013–1030. 10.2165/00023210-200317140-00002

11. Becker, H. C., & Mulholland, P. J. (2014). Neurochemical mechanisms of alcohol withdrawal. Handbook of clinical neurology, 125, 133–156. 10.1016/B978-0-444-62619-6.00009-4

12. Goethe, E. A., Deneen, B., Noebels, J., & Rao, G. (2023). The Role of Hyperexcitability in Gliomagenesis. International journal of molecular sciences, 24(1), 749. 10.3390/ijms24010749

13. Shalaby M, Moyer E, Buell KG, Bernard K, Gottlieb M. Presentations to United States emergency departments for gastroparesis, cyclic vomiting, and cannabinoid hyperemesis syndrome from 2016 to 2024. Am J Emerg Med. 2025;96:201–207. doi:10.1016/j.ajem.2025.06.067

14. Fiest KM, Sauro KM, Wiebe S, Patten SB, Kwon CS, Dykeman J, Pringsheim T, Lorenzetti DL, Jetté N. Prevalence and incidence of epilepsy: a systematic review and meta-analysis of international studies. Neurology. 2017;88(3):296–303. doi:10.1212/WNL.0000000000003509. PMID:27986877; PMCID:PMC5272794

15. Bakken IJ, Revdal E, Nesvåg R, Brenner E, Knudsen GP, Surén P, Ghaderi S, Gunnes N, Magnus P, Reichborn-Kjennerud T, Stoltenberg C, Trogstad LI, Håberg SE, Brodtkorb E. Substance use disorders and psychotic disorders in epilepsy: a population-based registry study. Epilepsy Res. 2014;108(8):1435–1443. doi:10.1016/j.eplepsyres.2014.06.021. PMID:25062893

16. Calonge, Q., Hanin, A., Januel, E., Guinebretiere, O., Nedelec, T., Le Gac, F., Chavez, M., Tezenas du Montcel, S., & Navarro, V. (2025). Incidence, mortality, and management of status epilepticus from 2012 to 2022: An 11-year nationwide study. Epilepsia. Advance online publication. 10.1111/epi.18627

17. Mantel N, Haenszel W. Statistical aspects of the analysis of data from retrospective studies of disease. J Natl Cancer Inst. 1959.

18. Breslow NE, Day NE. Statistical Methods in Cancer Research, Vol. I: The Analysis of Case-Control Studies. IARC, 1980.

19. Ng, S. K., Hauser, W. A., Brust, J. C., & Susser, M. (1988). Alcohol consumption and withdrawal in new-onset seizures. The New England journal of medicine, 319(11), 666–673. 10.1056/NEJM198809153191102

20. Walsh, S., Donnan, J., Fortin, Y., Sikora, L., Morrissey, A., Collins, K., & MacDonald, D. (2017). A systematic review of the risks factors associated with the onset and natural progression of epilepsy. Neurotoxicology, 61, 64–77. 10.1016/j.neuro.2016.03.011

21. Xiao Y, Nie Z, Huang J, Zhao J, Dong C, Zou Y, Li Z, Yan B, Hu Y, Yang F, Lee JW, Lin AP, Tobochnik S, Zhou M, Lei Z. Risk Factors and Prognostic Implications of Tumor-Related Epilepsy in Diffuse Glioma Patients: A Real-World Multicenter Study. Brain Behav. 2025 May;15(5):e70510. doi: 10.1002/brb3.70510. PMID: 40320911; PMCID: PMC12050638.

