## supplementarytables for "Alcohol use disorder and epileptogenesis in primary malignant brain tumors: temporal and tumor grade associations in a nationwide EHR cohort study"

Supplementary Figures

| Demographic Factor | Subgroup | % Seizure in Alcohol Users (Count) | % Seizure in Non-Alcohol Users (Count) | Odds Ratio (Alcohol vs Non-Alcohol) | 95% Confidence Interval | p-value |
| --- | --- | --- | --- | --- | --- | --- |
| **Age** | 18-40 | 60.8% (1,170 / 1,923) | 45.1% (20,992 / 46,555) | 1.89 | [1.72, 2.08] | <0.0001 |
|  | 40-65 | 56.8% (4,061 / 7,151) | 46.2% (48,998 / 105,999) | 1.50 | [1.43, 1.58] | <0.0001 |
|  | 65+ | 45.9% (2,010 / 4,378) | 37.2% (33,003 / 88,649) | 1.43 | [1.35, 1.52] | <0.0001 |
| **Sex** | Male | 52.3% (4,583 / 8,769) | 36.8% (44,537 / 121,869) | 1.88 | [1.80, 1.96] | <0.0001 |
|  | Female | 47.2% (1,793 / 3,796) | 33.4% (37,213 / 111,490) | 1.79 | [1.67, 1.91] | <0.0001 |
| **Race/Ethnicity** | White | 7.3% (5,354 / 73,339) | 4.2% (5,333 / 127,787) | 1.81 | [1.74, 1.88] | <0.0001 |
|  | Black or African Amer | 8.9% (827 / 9,278) | 5.2% (668 / 12,877) | 1.79 | [1.61, 1.99] | <0.0001 |
|  | Asian | 3.0% (72 / 2,387) | 1.3% (59 / 4,626) | 2.41 | [1.70, 3.41] | <0.0001 |
|  | AI/AN + NH/PI | 8.4% (95 / 1,127) | 3.9% (72 / 1,824) | 2.24 | [1.63, 3.07] | <0.0001 |
|  | Other | 6.4% (572 / 8,962) | 3.1% (450 / 14,686) | 2.16 | [1.90, 2.45] | <0.0001 |

Supplementary Table 1: Seizure prevalence in alcohol and non-alcohol users in count and percentage of population. Odds ratio of alcohol compared to non-alcohol users, with 95% CI and p-value (<0.05).

| **Time (months)** | **Alc→Sz** | **Sz→Alc** | **OR** | **95% CI (Lower-Upper)** | **p-value** |
| --- | --- | --- | --- | --- | --- |
| 0-6 | 447 | 389 | 1.16 | 1.01-1.34 | 0.041 |
| 6-12 | 188 | 161 | 1.17 | 0.95-1.45 | 0.158 |
| 12-18 | 163 | 112 | 1.47 | 1.15-1.87 | 0.002 |
| 18-24 | 110 | 94 | 1.17 | 0.89-1.55 | 0.290 |
| 24-30 | 106 | 93 | 1.14 | 0.86-1.51 | 0.391 |
| 30-36 | 89 | 51 | 1.76 | 1.24-2.48 | 0.002 |
| 36-42 | 69 | 56 | 1.23 | 0.87-1.76 | 0.281 |
| 42-48 | 62 | 50 | 1.24 | 0.85-1.81 | 0.296 |
| 48-54 | 45 | 41 | 1.10 | 0.72-1.68 | 0.745 |
| 54-60 | 43 | 33 | 1.31 | 0.83-2.06 | 0.300 |
| 60-66 | 52 | 34 | 1.53 | 0.99-2.37 | 0.066 |
| 66-72 | 40 | 23 | 1.74 | 1.04-2.92 | 0.043 |
| 72-78 | 29 | 21 | 1.38 | 0.79-2.43 | 0.321 |
| 78-84 | 28 | 21 | 1.33 | 0.76-2.35 | 0.390 |
| 84-90 | 18 | 11 | 1.64 | 0.77-3.47 | 0.265 |
| 90-96 | 23 | 14 | 1.65 | 0.85-3.20 | 0.188 |
| 96-102 | 13 | 12 | 1.08 | 0.49-2.38 | 1.000 |

Supplementary Table 2: Odds ratio of seizure/epilepsy onset after alcohol use diagnosis compared to alcohol use onset after seizure/epilepsy diagnosis (p < 0.05).
